# Private sector tuberculosis care quality during the COVID-19 pandemic: A repeated cross-sectional standardized patients study of adherence to national TB guidelines in urban Nigeria

**DOI:** 10.1101/2024.02.17.24302708

**Authors:** Angelina Sassi, Lauren Rosapep, Bolanle Olusola-Faleye, Elaine Baruwa, Ben Johns, Mohammad Abdullah Heel Kafi, Lavanya Huria, Nathaly Aguilera Vasquez, Benjamin Daniels, Jishnu Das, Chukwuma Anyaike, Obioma Chijioke-Akaniro, Madhukar Pai, Charity Oga-Omenka

## Abstract

Only a third of TB cases in Nigeria in 2020 were diagnosed and notified, in part due to low detection and underreporting from the private health sector. Using a standardized patient (SP) survey approach, we assessed how management of presumptive TB in the private sector aligns with national guidelines and whether this differed from a study conducted before the start of the COVID-19 pandemic. Thirteen standardized patients presented a presumptive TB case to 511 private providers in urban areas of Lagos and Kano states in May and June 2021. Private provider case management was compared with national guidelines divided into three main steps: SP questioned about cough duration; sputum collection attempted for TB testing; and non-prescription of anti-TB medications, antibiotics, and steroids. SP visits conducted in May-June 2021 were directly compared to SP visits conducted in the same areas in June-July 2019.

Overall, only 145 of 511 (28%, 95% CI: 24.5–32.5%) interactions were correctly managed according to Nigerian guidelines, as few providers completed all three necessary steps. Providers in 71% of visits asked about cough duration (362 of 511, 95% CI: 66.7–74.7%), 35% tested or recommended a sputum test (181 of 511, 95% CI: 31.3–39.8%), and 79% avoided prescribing or dispensing unnecessary medications (406 of 511, 95% CI: 75.6–82.8%). COVID-19 related questions were asked in only 2.4% (12 of 511, 95% CI: 1.3–4.2%) of visits. During the COVID-19 pandemic, few providers completed all steps of the national guidelines. Providers performed better on individual steps, particularly asking about symptoms and avoiding prescription of harmful medications. Comparing visits conducted before and during the COVID-19 pandemic showed that COVID-19 did not significantly change the quality of TB care.

**Key Messages:** *What is already known on this topic:*

- Less than half of new TB cases in Nigeria are diagnosed and notified. As most initial health care seeking for TB in Nigeria occurs in the private sector, increasing the quality of TB care in the private sector is of great importance.
- COVID-19 may have put further stressors on TB care quality due to changes in care seeking behavior, stigma against COVID-19, and disproportionate attention at the health system level on pandemic control.
- This study explored whether private providers’ practices are in alignment with national standards for TB screening in Nigeria, how these practices have changed following the onset of the COVID-19 pandemic, and what factors are associated with providers that deliver clinically correct TB screening services.

*What this study adds:*

- Fewer than one-third of the SP visits conducted in this study were correctly managed according to the Nigerian National TB and Leprosy Control Program guidelines.
- Clinical correctness of TB care in the private sector of urban Nigeria has not been majorly affected by COVID-19 according to our study results.
- Our results indicate that very little observed attention was paid to COVID-19 in this sample of private facilities.

*How this study might affect research, practice or policy:*

- Increased efforts to engage and support private providers, and implementing solutions such as working with drug shop proprietors to make referring for testing a standard part of their practice may help reduce the testing bottleneck at drug shops.
- Although Nigeria has maintained pre-pandemic levels of TB notification, it is important to establish high-quality screening by all providers to find the missing patients with TB and close the gap in TB notification.

## Introduction

Nigeria is one of eight countries that account for more than two-thirds of new TB cases each year according to WHO estimates (1). There were 467,000 new TB cases in Nigeria in 2021, but only 44% of those cases were diagnosed and notified (1). Though there remains a gap between new cases and TB notifications, Nigeria has reported increased TB notifications each year since 2018 (2).

One of the major challenges is that many providers working in Nigeria’s large private healthcare sector are not adequately linked to Nigeria’s National TB and Leprosy Control Program (NTBLCP) activities. Between 66 and 92% of initial care-seeking for respiratory diseases in Nigeria occurs in the private sector (3–5), but the private sector accounted for just 12% of TB notifications in 2018 (5). The Federal Ministry of Health has committed to engaging and strengthening the private sector to scale up sustainable case finding, treatment, and notification of TB in Nigeria (6,7); in 2020, the share of private sector TB notifications increased to 26% in 2020 (8). Ensuring timely and accurate TB case detection in the private sector is crucial to understanding the quality gaps in TB care among this group of providers (9,10).

The use of standardized patients (SPs) – individuals recruited from the local community to present the same case to multiple providers in a blinded fashion – is considered the gold standard for measurement of health care quality and has been used increasingly in LMICs (11). A standardized patients (SP) study conducted in 2019 among 837 private facilities and 206 public providers in urban Lagos and Kano showed that while more than 70% of providers correctly screened for TB, a minority of providers met the criteria for correct management – defined as screening, recommended testing, and refraining from prescribing or dispensing inappropriate medications – of SPs presenting with “textbook” TB symptoms, i.e. three weeks of cough, mild, fever, and some weight loss (12).

Globally, COVID-19-related health care disruptions have reversed years of progress in strengthening TB care (1,13). In many countries, lockdowns, movement restrictions, and fears of acquiring COVID-19 at health facilities greatly limited patient care seeking, while facility closures and provider priorities shifting to COVID-19 further constrained available TB services on the supply side (14,15). Nigeria is one of a small group of high-TB burden countries that did not experience a reduction in TB notifications between 2020 and 2022 (1,13). This may be due in part to the relatively low COVID-19 testing rates and/or low caseload in sub-Saharan African countries including Nigeria (16–18), though serological studies indicate available figures may underestimate COVID-19 cases (19–21). Additionally, efforts by the public health system to integrate TB active case finding into COVID-19 sensitization in all states may have contributed to the stability of TB notifications in Nigeria (2).

Although Nigeria may have weathered the pandemic differently than other countries, COVID-19 may have put further stressors on TB care quality due to changes in care seeking behavior, stigma against COVID-19, and disproportionate attention at the health system level on pandemic control (14,22,23). Additionally, the overlap in symptoms between COVID-19 and TB could be hypothesized to increase misdiagnosis of TB cases as COVID-19 cases or vice-versa (24,25). As part of a multi-country research effort to examine how COVID-19 has affected the delivery of private sector TB services in high-burden countries (26–29), this study explored whether private providers’ practices are in alignment with national standards for TB screening in Nigeria, how these practices have changed following the onset of the COVID-19 pandemic, and what factors are associated with providers that deliver clinically correct TB screening services.

## Methods

### Study setting

This cross-sectional SP study was conducted in urban areas in two of Nigeria’s most populated states, Lagos and Kano, which have a combined population of over 24 million people (30). Between 2018 and 2021, both of these states received support from the United States Agency for International Development (USAID)-funded Sustaining Health Outcomes through the Private Sector (SHOPS) Plus program to implement a public-private mix (PPM) approach to improve private sector TB case detection and treatment (12). In both states, the SHOPS Plus network worked with four different types of health facilities: clinical facilities, stand-alone private laboratories, private community pharmacies, and medicine vendors. These providers were trained in TB screening, diagnosis, and treatment standards, then organized into “hub-and-spoke” clusters, with laboratories, pharmacies, and drug shops serving as “spokes” to drive patient traffic into clinical facility hubs for further assessment and, where necessary, treatment. Other details about this setting are included in the prior study (12).

As of 11 February 2024, there have been 267,173 reported COVID-19 cases and 3,155 confirmed deaths in Nigeria (31). The Government of Nigeria issued a series of movement restrictions in several states beginning on March 30^th^, 2020, which began to ease after five weeks (32). In the early months of the COVID-19 pandemic, only public facilities in Nigeria were allowed to treat COVID-19 patients and nationwide screening capacity was limited. In July 2020, the Federal Ministry of Health announced that COVID-19 sample collection would be scaled up to all eligible public and private hospitals in Nigeria (32,33). Evidence from a mapping survey of private healthcare facilities in Lagos and Kano, Nigeria indicates that COVID-19 had a temporary impact on private sector TB care, lasting for the first three to six months of 2020 (26).

### Sampling frame and sample size

This study aimed to assess clinical correctness of TB care within the same study area and sampling frame as was reported in Rosapep et al., 2022 (12). While the original study measured quality of TB care among private facilities within and outside of the SHOPS Plus network, we opted to narrow our focus on clinical facilities and drug shops as we hypothesized these facility types may be particularly susceptible to changes in patient load during the COVID-19 pandemic. All 357 private clinical facilities from the 2019 survey were included in the sampling frame. To create the drug shop sampling frame, we randomly selected 200 drug shops (100 in each state) for verification of operating status. The team conducted verifications based on what could be observed outdoors to avoid having data collectors enter facilities. The verification occurred between 23-29 March 2021. Out of 200 randomly selected drug shops, 171 were verified as being in operation. 143 facilities (72 in Kano and 71 in Lagos) were randomly selected from the list of operating drug shops. The remainder were designated as potential replacement facilities for those found to be non-operational during field work.

### Data collection

SP interactions occurred primarily from 3-17 May 2021, with five other interactions completed in June 2021 in facilities that were closed during the main fieldwork period. All visits were unannounced and conducted using a “textbook” case of presumptive TB, which describes a patient with two to three weeks of cough with sputum, mild fever, and some weight loss and loss of appetite (see Table 1) (11,12). This case presentation should prompt providers to screen for TB and initiate further diagnostic testing (i.e., GeneXpert MTB/RIF (Xpert) (Cepheid, Sunnyvale, CA), Acid-Fast Bacilli (AFB) microscopy, culture, Drug susceptibility testing (DST) and/or chest X-ray (CXR)). After each interaction, SPs were debriefed by field supervisors using a standardized structured exit questionnaire deployed on SurveyCTO (Dobility, Inc.). Questions were added to this survey that sought to understand the extent to which providers asked patients about COVID-19, screened, or tested them for COVID-19, or suspected COVID-19 as their diagnosis.

**Table 1:** Description of standardized patient scenario case presentation.

| Case | Type of facilities | Presenting condition or opening statement by SP | Expected management |
| --- | --- | --- | --- |
| Case 1:<br>presumptive TB<br>patient scenario | Private SHOPS Plus<br>facilities (clinics, drug<br>shops)<br>Private non-SHOPS<br>Plus clinical facilities | *“I am having fever and*<br>*cough that is not getting*<br>*better.”* | Provider expected to:<br>• ask more questions about symptoms and duration<br>• initiate further diagnostic testing (sputum sample, CXR, or referring patient elsewhere if the facility is not testing for TB)<br>• refrain from dispensing inappropriate (i.e., anti-TB drugs, fluoroquinolones, steroids) or unnecessary (i.e., other antibiotics) medications. |

### Data analysis

Our main outcome of interest was binary correct case management, based on NTPLCP guidelines (34), using the same thresholds used in the previous SP study (12), to allow comparability between the pre-and during-pandemic data. Clinical providers must meet all three criteria to demonstrate correct case management: 1) confirmation of core TB symptoms, referred to as the “screening step”, defined as asking about productive cough lasting two weeks or longer and at least one of the following symptoms: fever, blood in sputum, chest pain, unexplained weight loss, difficulty breathing, or night sweats; 2) recommendation or attempt to take any appropriate diagnostic (CXR or sputum sample taken or attempted) or provision of a referral to another public or private clinical facility for testing, referred to as the “testing step”; 3) refrain from prescribing or dispensing antibiotics (including anti-TB drugs and fluoroquinolones) and steroids. The standards for drug shop providers are the same for all three criteria, except that drug shop providers need only confirm a cough of two weeks or longer to meet criteria 1 (confirmation of core TB symptoms). To match the previous study, drug shop providers were not expected to order tests or dispense medications such as anti-TB drugs, antibiotics, or steroids, though they were expected to attempt to collect or refer patients to clinical facilities for diagnostic tests (12).

We calculated proportions and 95% confidence intervals (CI) for all the component elements described above and the composite binary measure of correct management. We compared data from SP visits conducted in 2021 with those conducted in the 2019 study within the same sampling area using logistic regression. We used a generalized estimating equation (GEE) model to assess facility, provider, and visit characteristics associated with correct case management considering the time of each study (pre-COVID-19 and during-COVID-19). We used GEE models to account for the violation of independence in our data due to our design, i.e., correct case management by the same provider over time are expected to be more similar than scores between different providers. Potential covariates were selected based on prior literature and observed characteristics of the interaction: categorical variables representing year of survey (2019 and 2021), facility location (state), length of the visit in minutes, type of facility visited, and gender of the most senior provider that attended to the SP (highest provider gender). Due to the large difference in support and training provided to SHOPS Plus-supported clinical facilities compared to those clinical facilities outside the SHOPS Plus network (referred to as “non-network clinical facilities”), we regarded SHOPS Plus clinical facilities and non-network clinical facilities as distinct facility types, with SHOPS Plus supported drug shops as a third facility type (there were no non-network drug shops included in this study nor the 2019 SP study). These models were computed for the overall binary correct management outcome measure as well as for the three individual component elements in the correct management measure. Regression results are reported as simple odds ratios (ORs) and adjusted odds ratios (aORs) with 95% confidence intervals (CI) for each variable.

### Ethical considerations

This study received ethical approval from the ethics boards of the authors’ institutes and from the Health Research Ethics Committee (HREC) in the two Nigerian states: HREC Lagos State University Teaching Hospital (LASUTH) (LREC/06/10/1517) and HREC Kano State MoH (MOH/Off797/T.I/2168).

## Results

SPs completed 511 interactions out of 543 attempted for a total response rate of 94.11%. All SPs were able to complete presentation of their case and no SPs reported any errors or provider detections. All SP interactions occurred in May and June 2021, and during this period, reported COVID-19 numbers in Lagos and Kano were low and there was no new surge or wave (35).

### Description of the standardized patient visits

Table 2 presents descriptive statistics of the SP visits conducted in 2019 and 2021 disaggregated by study year and facility type (non-network clinical facilities, SHOPS Plus-supported clinical facilities, and SHOPS Plus-supported drug shops). The remainder of this section will concentrate on the description of the 511 SP visits conducted in 2021. Seven out of 13 SPs were male. SPs ranged in age between 23 and 35 years old, with a mean age of 30 (Standard Deviation (SD) = 4.26). About half of visits were conducted by female SPs (51%, 258/511). The average visit length across all facilities was 29 minutes (SD=27). SPs were examined or asked about their condition by one person in 96% of visits made to drug shops (172/179). The highest-ranking provider who examined the SP or asked about their condition was a physician in 85% of SHOPS Plus clinical facility visits (228/268) and 77% of non-network clinical facility visits (49/64), and a pharmacist or drug shop purveyor in 93% of drug shop visits (167/179). In 73% of all SP visits, the highest-ranking provider was male (374/511).

**Table 2:**
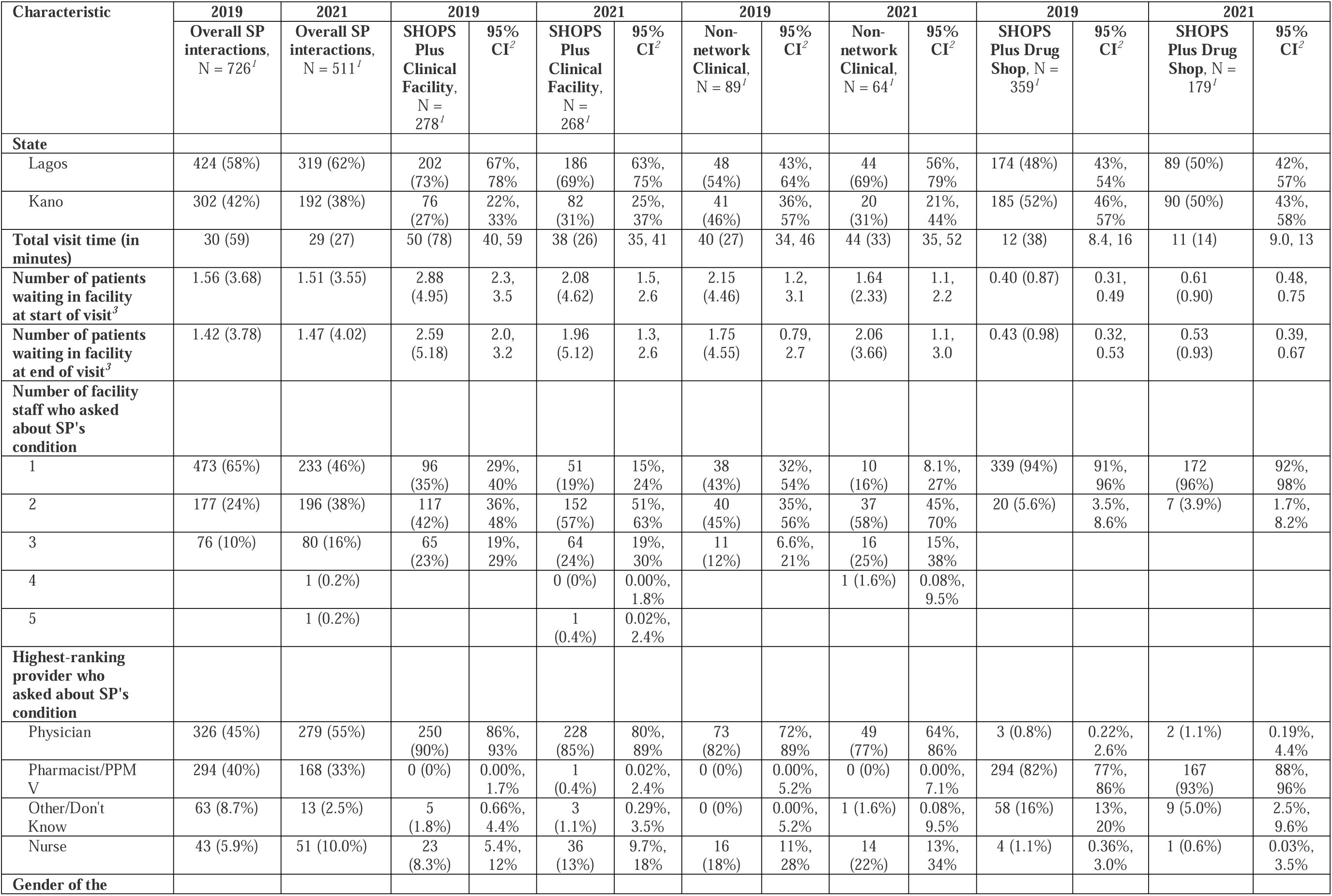
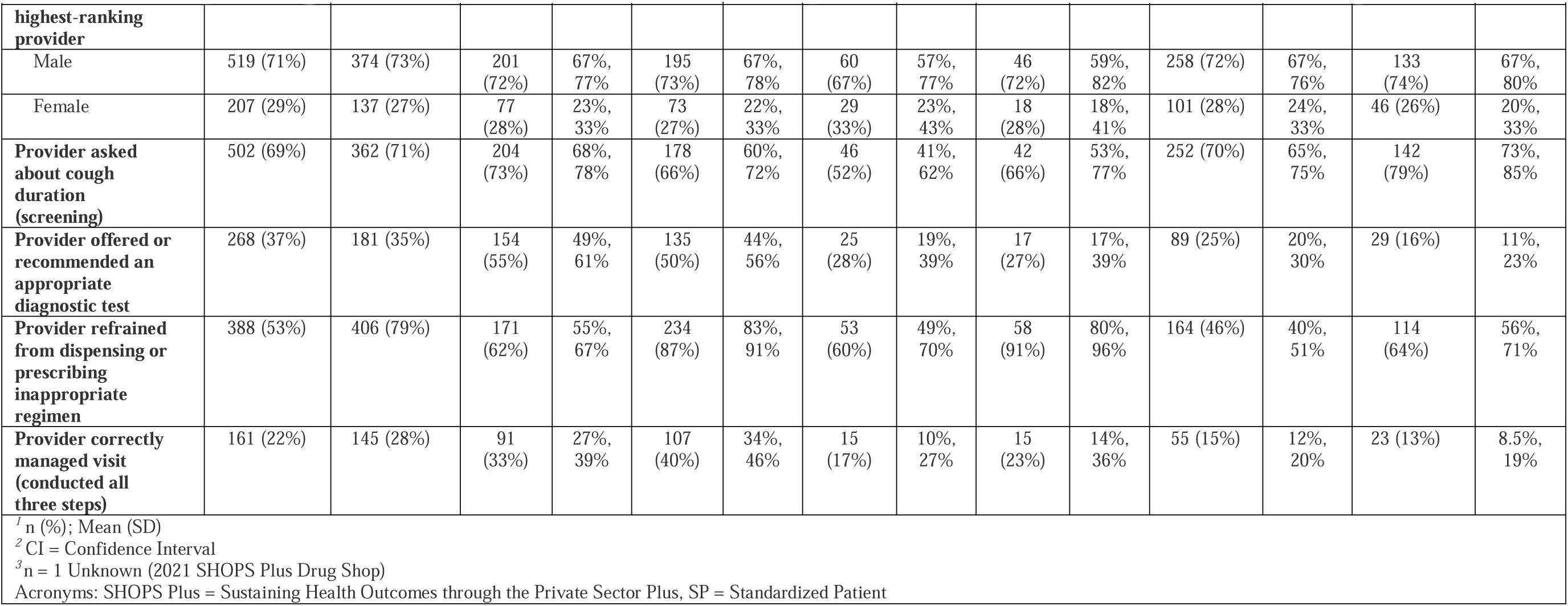
Descriptive statistics of standardized patient visits in both study years.

Overall, 28% of all SP visits were correctly managed across all three steps of the NTBLCP guidelines (145/511, 95% CI: 24.5–32.5%). Correct management was observed more in SP visits made to SHOPS Plus clinical facilities (40%, 107/268, 95% CI: 34.0–46.1%) compared to non-network clinical facilities (15/64, 23%, 95% CI: 13.8–35.7%) and SHOPS Plus-network drug shops (23/179, 13%, 95% CI: 8.3–18.7%).

Providers were comparatively more successful in completing each individual step of correct case management (Figure 1). Providers successfully confirmed TB symptoms in 71% of all visits (362/511, 95% CI: 66.7–74.7%). Over two-thirds of providers in each facility type — 66% of providers in SHOPS Plus clinical facilities (178/268, 95% CI: 66.4–72.0%), 66% of providers in non-network clinical facilities (42/64, 95% CI: 52.6–76.8%), and 79% of providers in drug shops (142/179, 95% CI: 72.5–84.9%) — completed the screening step.

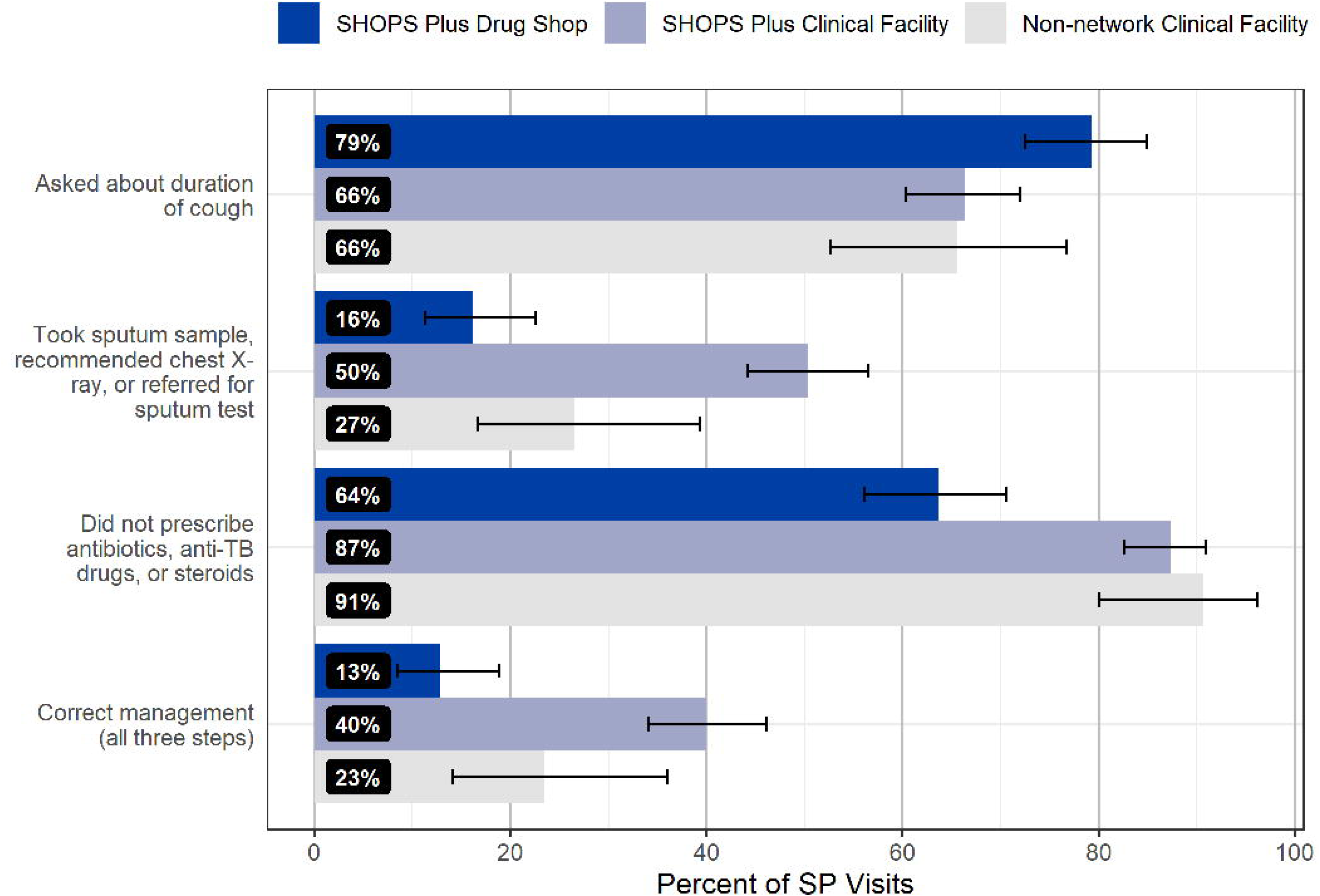

Providers offered or recommended an appropriate diagnostic test in just over one third (35%) of visits (181/511, 95% CI: 31.3–39.8%). Any type of lab or diagnostic tests was offered or recommended by providers in 68.8% of non-network clinical facility visits (44/64, 95% CI: 55.8–79.4%), 80.2% of SHOPS-Plus supported clinical facility visits (215/268, 95% CI: 74.8–84.7%), and 20.1% of drug shop visits (36/64, 95% CI: 14.7–26.9%) (Figure 2). SPs were offered or recommend any type of sputum TB test in 23.1% of all visits (118/511, 95% CI: 19.6– 27.0%). While the most common TB tests offered were acid-fast bacillus (AFB) and GeneXpert (AFB: 11.7% [60/511], 95% CI: 9.1–14.9%; Xpert: 11.0% [56/511], 95% CI: 8.4–14.1%), the most common tests offered or recommended overall were for malaria and typhoid (malaria: 32.5% [166/511], 95% CI: 28.5–36.8%; typhoid: 20.9% [107/511], 95% CI: 17.5–24.8%).

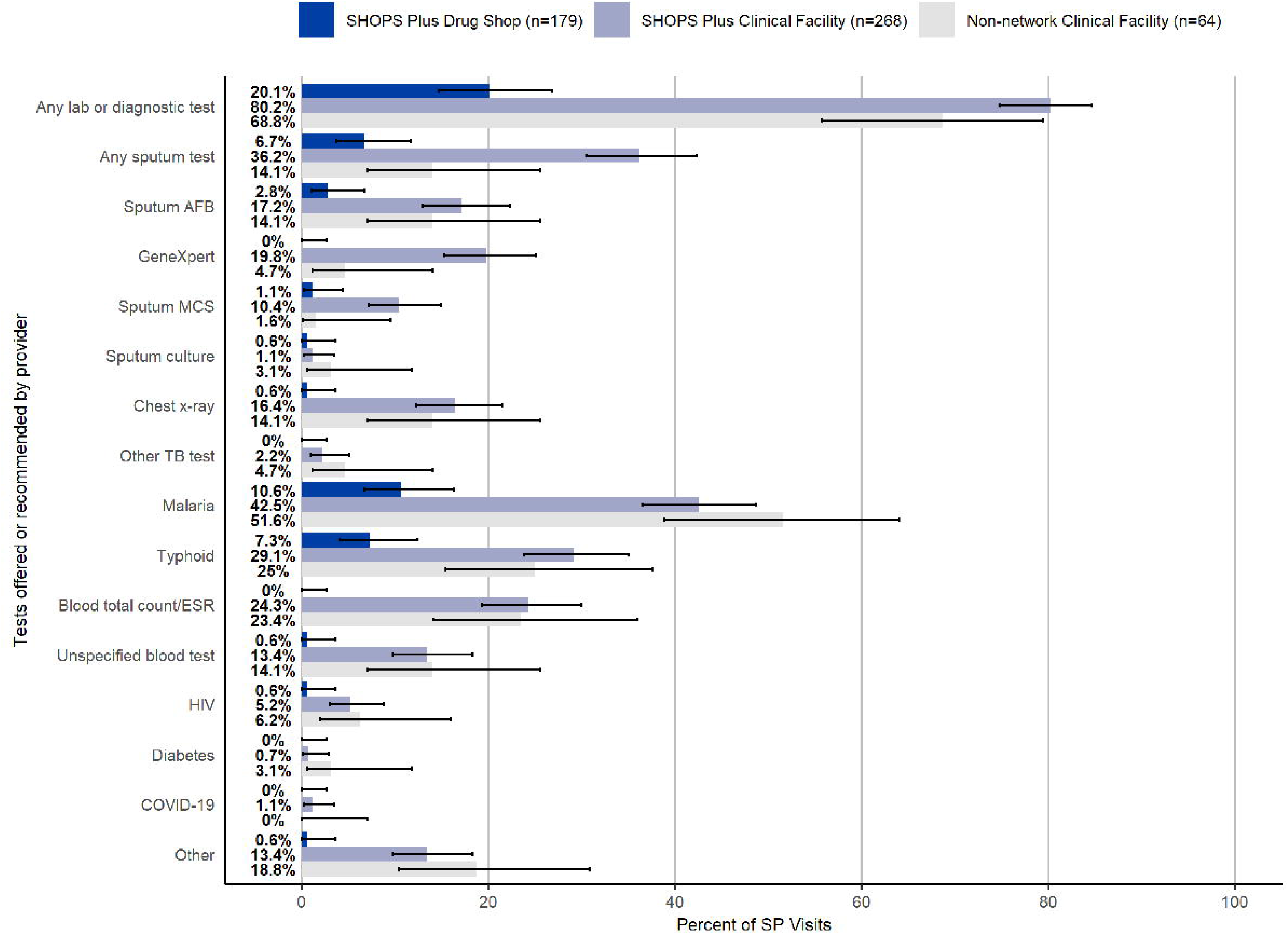

Providers in 79% of visits successfully refrained from dispensing or prescribing antibiotics, TB medication, or steroids (406/511, 95% CI: 75.6–82.8%). One or more medications of any kind were prescribed or dispensed in 27% of all SP visits (138/511, 95% CI: 23.2–31.1%). No anti-tuberculosis medication was prescribed or dispensed in any SP visit. Other drugs considered to be inappropriate to administer to people with unconfirmed TB infection because they can delay TB detection – fluoroquinolones, or steroids – were dispensed or prescribed in less than 5% of SP visits (fluoroquinolones: 4.3% [22/511], 95% CI: 2.8–6.5%; steroids: 0.8% [4/511], 95% CI: 0.3–2.1%). Other non-anti-TB antibiotics were the most prescribed or dispensed type of medication, in 17.6% of all SP visits. (90/511, 95% CI: 14.5– 21.3%). Providers at non-network clinical facilities and SHOPS Plus-supported clinical facilities were less likely to prescribe or dispense one or more medications compared to providers at SHOPS Plus supported drug shops (non-network clinical facilities: 12.5% [8/64], 95% CI: 5.9– 23.7%; SHOPS Plus clinical facilities: 16.4% [44/268] 95% CI: 12.3–21.5%; drug shops: 48% [86/179], 95% CI: 40.6%–55.6%) (Figure 3).

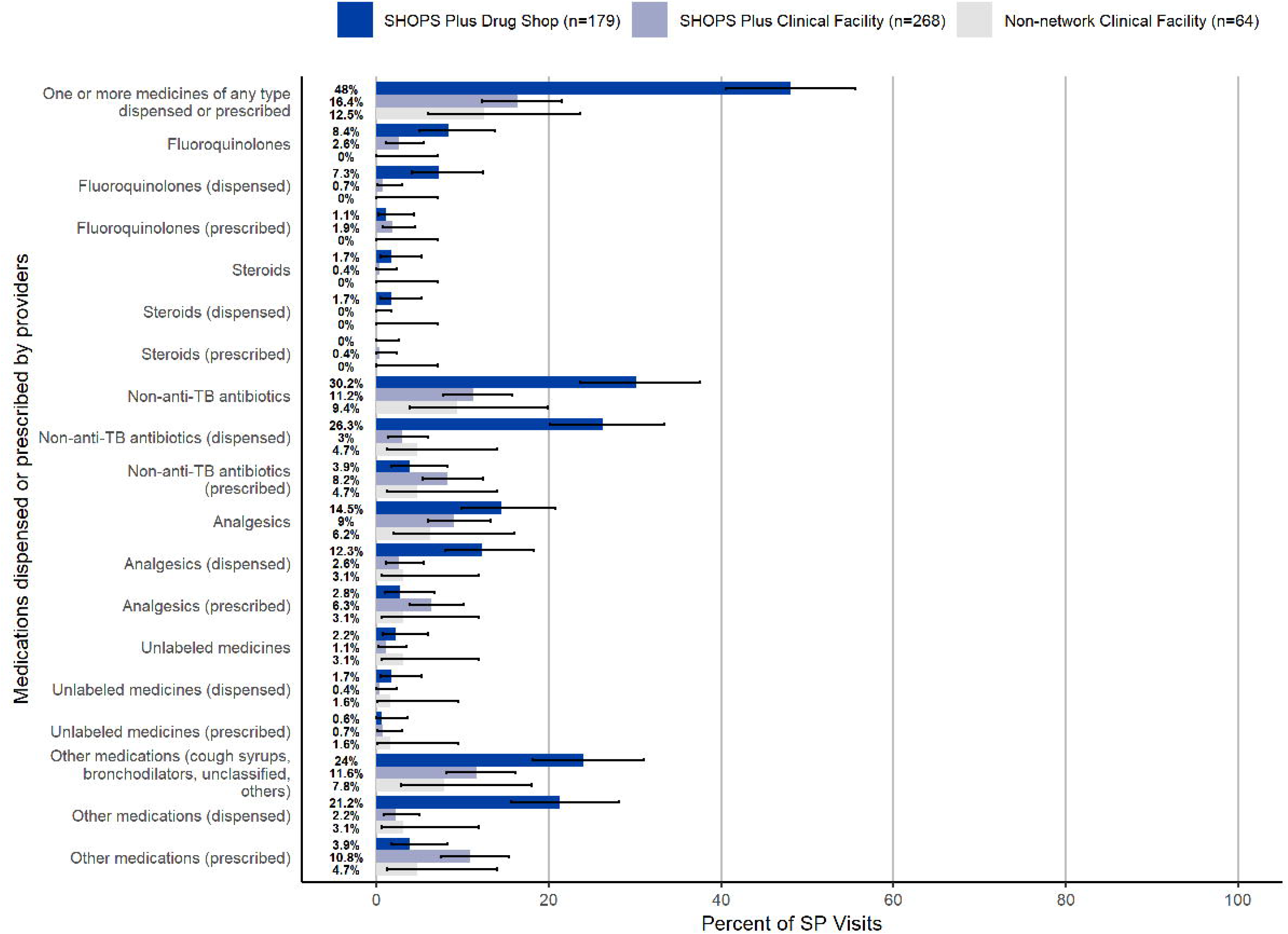

COVID-19 was mentioned in 12 of the 511 SP visits (2.4%, 95% CI: 1.3–4.2%). SPs in three of the 511 visits were asked if they had ever had COVID-19. SPs in six of the 511 visits were asked if they had been with anyone known to have COVID-19 or have been suspected to have had COVID-19. SPs in three of the 511 visits were offered COVID-19 tests. The provider mentioned a suspicion of COVID-19 during the conversation in five SP visits.

### Regression comparison of pre-COVID-19 and during-COVID-19 standardized patient study results

Results from the regression analysis can be found in Tables 3 and 4 below. There was no significant difference across the two study years in overall correct case management while adjusting for state, length of visit, facility type and network status, and highest provider gender [adjusted OR (aOR): 1.21, 95% CI: 0.92–1.58, p=0.2]. Compared to SP visits conducted in Lagos state, Kano providers were less likely to correctly manage an SP, holding all other variables constant (aOR: 0.50, 95% CI: 0.37–0.68, p<0.001). SPs who visited non-network clinical facilities or SHOPS Plus supported drug shops were significantly less likely to be correctly managed compared to SHOPS Plus supported clinical facilities (non-network clinical facility: aOR = 0.46, 95% CI = 0.29–0.71, p<0.001; SHOPS Plus drug shop: aOR = 0.36, 95% CI = 0.26–0.49, p<0.001). No significant differences in overall correct management were observed among visits with an increase of 10 minutes in visit length (aOR: 1.01, 95% CI: 0.99– 1.04, p=0.4).

**Table 3:**
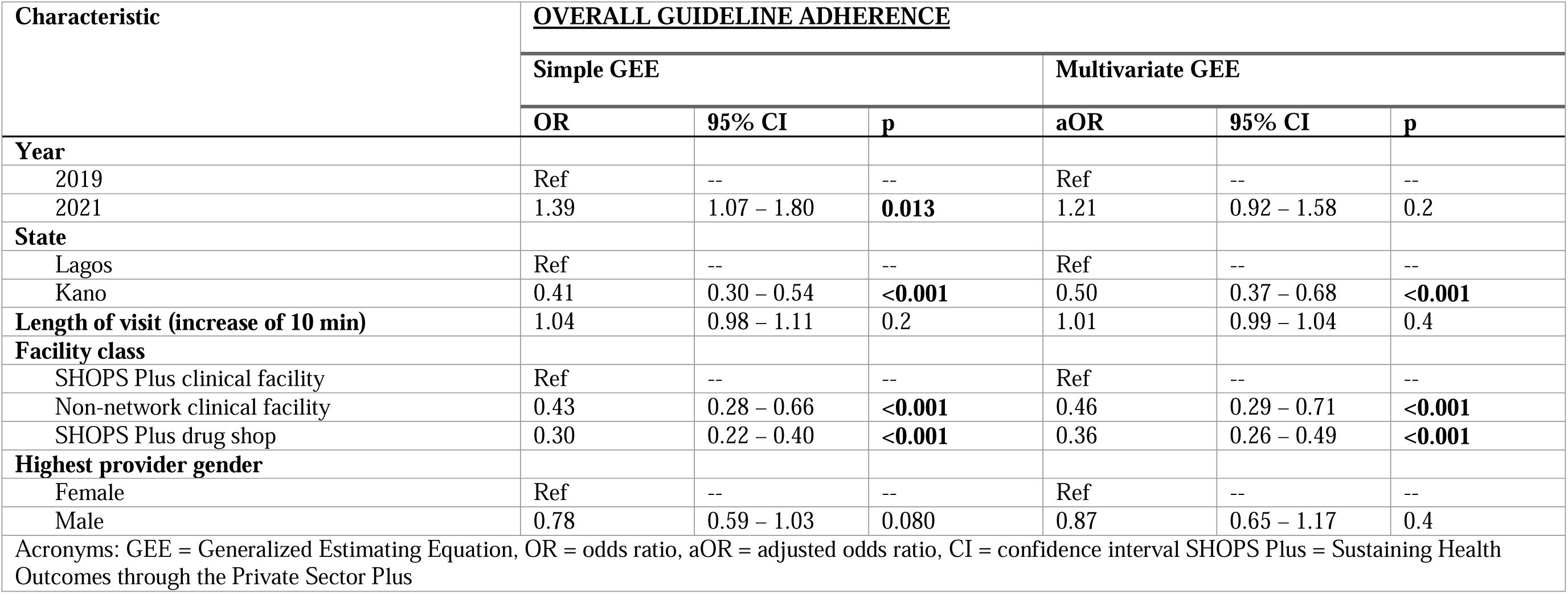
Simple and multivariate regression results on overall guideline adherence.

| Characteristic | **<u>OVERALL GUIDELINE ADHERENCE</u>** |  |  |  |  |  |
| --- | --- | --- | --- | --- | --- | --- |
|  | **Simple GEE** |  |  | **Multivariate GEE** |  |  |
|  | **OR** | **95% CI** | **p** | **aOR** | **95% CI** | **p** |
| **Year** |  |  |  |  |  |  |
| 2019 | Ref | -- | -- | Ref | -- | -- |
| 2021 | 1.39 | 1.07 – 1.80 | **0.013** | 1.21 | 0.92 – 1.58 | 0.2 |
| **State** |  |  |  |  |  |  |
| Lagos | Ref | -- | -- | Ref | -- | -- |
| Kano | 0.41 | 0.30 – 0.54 | **<0.001** | 0.50 | 0.37 – 0.68 | **<0.001** |
| **Length of visit (increase of 10 min)** | 1.04 | 0.98 – 1.11 | 0.2 | 1.01 | 0.99 – 1.04 | 0.4 |
| **Facility class** |  |  |  |  |  |  |
| SHOPS Plus clinical facility | Ref | -- | -- | Ref | -- | -- |
| Non-network clinical facility | 0.43 | 0.28 – 0.66 | **<0.001** | 0.46 | 0.29 – 0.71 | **<0.001** |
| SHOPS Plus drug shop | 0.30 | 0.22 – 0.40 | **<0.001** | 0.36 | 0.26 – 0.49 | **<0.001** |
| **Highest provider gender** |  |  |  |  |  |  |
| Female | Ref | -- | -- | Ref | -- | -- |
| Male | 0.78 | 0.59 – 1.03 | 0.080 | 0.87 | 0.65 – 1.17 | 0.4 |
| Acronyms: GEE = Generalized Estimating Equation, OR = odds ratio, aOR = adjusted odds ratio, CI = confidence interval SHOPS Plus = Sustaining Health Outcomes through the Private Sector Plus |  |  |  |  |  |  |

**Table 4:** Simple and multivariate regression results on each step of the correct management guideline.

|  | <u>STEP 1 - SCREENING</u> |  |  |  |  |  | <u>STEP 2 - TESTING</u> |  |  |  |  |  | <u>STEP 3 – AVOIDING INAPPROPRIATE PRESCRIBING</u> |  |  |  |  |  |
| --- | --- | --- | --- | --- | --- | --- | --- | --- | --- | --- | --- | --- | --- | --- | --- | --- | --- | --- |
| Characteristic | Simple GEE |  |  | Multivariate GEE |  |  | Simple GEE |  |  | Multivariate GEE |  |  | Simple GEE |  |  | Multivariate GEE |  |  |
|  | OR | 95% CI | p | aOR | 95% CI | p | OR | 95% CI | p | aOR | 95% CI | p | OR | 95% CI | p | aOR | 95% CI | p |
| **Year** |  |  |  |  |  |  |  |  |  |  |  |  |  |  |  |  |  |  |
| 2019 | Ref | -- | -- | Ref | -- | -- | Ref | -- | -- | Ref | -- | -- | Ref | -- | -- | Ref | -- | -- |
| 2021 | 1.08 | 0.85 – 1.39 | 0.5 | 1.12 | 0.88, 1.44 | 0.4 | 0.94 | 0.74 – 1.19 | 0.6 | 0.76 | 0.59 – 0.98 | **0.033** | 3.37 | 2.60 – 4.37 | **<0.001** | 3.10 | 2.39 – 4.02 | **<0.001** |
| **State** |  |  |  |  |  |  |  |  |  |  |  |  |  |  |  |  |  |  |
| Lagos | Ref | -- | -- | Ref | -- | -- | Ref | -- | -- | Ref | -- | -- | Ref | -- | -- | Ref | -- | -- |
| Kano | 0.85 | 0.67 – 1.09 | 0.2 | 0.75 | 0.55, 1.01 | 0.056 | 0.57 | 0.45 – 0.73 | **<0.001** | 0.77 | 0.59 – 1.01 | 0.057 | 0.56 | 0.44 – 0.71 | **<0.001** | 0.64 | 0.49 – 0.84 | **0.001** |
| **Length of visit (increase of 10 min)** | 1.04 | 0.95 – 1.13 | 0.4 | 1.09 | 0.90, 1.31 | 0.4 | 1.12 | 0.99 – 1.27 | 0.082 | 1.03 | 0.97 – 1.09 | 0.3 | 1.01 | 0.97 – 1.06 | 0.6 | 0.98 | 0.95 – 1.01 | 0.12 |
| **Facility class** |  |  |  |  |  |  |  |  |  |  |  |  |  |  |  |  |  |  |
| SHOPS Plus clinical facility | Ref | -- | -- | Ref | -- | -- | Ref | -- | -- | Ref | -- | -- | Ref | -- | -- | Ref | -- | -- |
| Non-network clinical facility | 0.58 | 0.40 – 0.84 | **0.004** | 0.60 | 0.42, 0.88 | **0.008** | 0.34 | 0.23 – 0.50 | **<0.001** | 0.33 | 0.22 – 0.50 | **<0.001** | 0.92 | 0.61 – 1.38 | 0.7 | 1.04 | 0.69 – 1.58 | 0.8 |
| SHOPS Plus drug shop | 1.17 | 0.90 – 1.53 | 0.2 | 1.62 | 0.93, 2.82 | 0.088 | 0.25 | 0.19 – 0.33 | **<0.001** | 0.27 | 0.20 – 0.37 | **<0.001** | 0.37 | 0.29 – 0.48 | **<0.001** | 0.43 | 0.32 – 0.57 | **<0.001** |
| **Highest provider gender** |  |  |  |  |  |  |  |  |  |  |  |  |  |  |  |  |  |  |
| Female | Ref | -- | -- | Ref | -- | -- | Ref | -- | -- | Ref | -- | -- | Ref | -- | -- | Ref | -- | -- |
| Male | 1.68 | 1.30 – 2.19 | **<0.001** | 1.80 | 1.36, 2.38 | **<0.001** | 0.77 | 0.60 – 1.00 | **0.046** | 0.79 | 0.60 – 1.04 | 0.10 | 0.88 | 0.68 – 1.14 | 0.3 | 0.97 | 0.73 – 1.29 | 0.8 |
| Acronyms: GEE = Generalized Estimating Equation, OR = odds ratio, aOR = adjusted odds ratio, CI = confidence interval SHOPS Plus = Sustaining Health Outcomes through the Private Sector Plus |  |  |  |  |  |  |  |  |  |  |  |  |  |  |  |  |  |  |

Multivariate regression on only the screening component of the correct management measure revealed that non-network clinical facilities were less likely to properly screen for TB symptoms compared to SHOPS Plus clinical facilities (aOR: 0.60, 95% CI: 0.42–0.88, p=0.008). Additionally, male providers were more likely to pass the screening step compared to female providers (aOR: 1.80, 95% CI: 1.36–2.38, p<0.001). No other significant associations were found between completion of the screening step and study year, state, length of visit, and facility type and network status.

For the testing step, visits conducted in 2021 were less likely to properly recommend SPs to testing compared to visits conducted in 2019 (aOR: 0.76, 95% CI: 0.59–0.98, p=0.033). Additionally, non-network clinical facilities and SHOPS Plus drug shops were less likely to properly screen for TB symptoms compared to SHOPS Plus clinical facilities (non-network clinical facilities: aOR=0.33, 95% CI: 0.22–0.50, p<0.001; SHOPS Plus drug shops: aOR=0.27, 95% CI: 0.20–0.37, p<0.001). No other significant associations were found between completion of the testing step and state, length of visit, and gender of the most senior provider who attended to the SP.

Providers in SP visits conducted in 2021 were more likely to avoid inappropriate prescribing compared to visits conducted in 2019 (aOR: 3.10, 95% CI: 2.39–4.02, p<0.001). Additionally, Kano facilities were less likely to avoid inappropriate prescribing compared to Lagos facilities (aOR: 0.64, 95% CI: 0.49–0.84, p=0.001). Finally, SHOPS Plus supported drug shops were less likely to avoid inappropriate prescribing compared to SHOPS Plus supported clinical facilities (aOR: 0.43, 95% CI: 0.32–0.57, p<0.001). No other significant associations were observed between the avoiding inappropriate prescribing step of the correct management guideline and length of visit, facility class and network status, and gender of the most senior provider who attended to the SP.

## Discussion

Nigeria is one of the highest TB burden countries in the world and has a large private health sector. Engaging and improving quality of TB care in the private sector is a key priority for the National TB Program. In our SP survey, we compared quality of TB care during the COVID-19 pandemic with prior, published data pre-pandemic in 2019.

As in the 2019 study, we applied a stringent three-criteria requirement for measuring correct management of standardized patients presenting a case of classic TB. Our findings showed that fewer than one-third of the SP visits conducted in this study were correctly managed according to all three components of the NTBLCP guidelines. This is suboptimal and strongly suggests the need for greater private provider engagement efforts to improve quality of TB care. Nevertheless, providers largely managed cases appropriately on the individual components of these guidelines, particularly the screening step and avoiding prescription of inappropriate medication. Providers in just over a third of visits offered or recommended an appropriate diagnostic test, which was the main barrier to guideline adherence observed in this study. The proportion of providers who offered or recommended an appropriate diagnostic test in SHOPS Plus-supported clinical facilities and in non-network clinical facilities was higher than that among drug shop proprietors. This result is expected in this setting as drug shops typically dispense medication and are not generally expected to refer their clients for testing. The SHOPS Plus program has supported drug shops to increase referrals for testing by providing facilities with sputum cups and access to sputum transportation, as well as offering incentives for participation in these efforts. Despite this training, drug shop providers may still be reluctant to divert from the behavior typically expected from their customers, i.e. dispensing medication in a quick fashion (36,37). The training and support provided by the SHOPS Plus program likely also explains the difference observed between SHOPS Plus clinical facilities and non-network clinical facilities in overall guideline adherence and completion of the screening and testing steps of the guideline. These results are a strong indication that trained facilities are more equipped to follow NTBLCP guidelines.

Our results also indicate that very little observed attention was paid to COVID-19 in this sample of private facilities. Given the overlap in symptoms between TB and COVID-19, we expected more providers to assume that SPs had COVID-19 and insistent on COVID-19 testing. This could be due to the transient impact COVID-19 had on tuberculosis services in Nigeria (26), limited COVID-19 service coverage allowed in the private sector (32), or limited availability of COVID-19 screening and diagnostic tools in Nigeria as a whole (38,39). It is also possible that the networked providers were able to correctly differentiate TB symptoms from COVID-19 symptoms once the SPs described their symptoms, since the SHOPS Plus program had trained networked providers on this distinction at the start of the COVID-19 pandemic.

A comparison of this study’s results with the SP study conducted in the same sampling area in 2019 revealed few major changes in overall correct management. Compared to the pre-COVID-19 SP study, providers in the during-COVID-19 study were less likely to properly recommend SPs for testing. This reduction in recommendations for testing could be caused by an increased reluctance by providers to recommend additional healthcare services to their clients because of the overall impact of COVID on TB care reported in numerous other studies (15,23,40). However, this explanation may not suffice in the Nigeria context due to the relatively small and immediate impact of COVID-19 restrictions on access to healthcare services. Therefore, a more likely explanation may be the negative effect of COVID-19 on the providers’ willingness to collect sputum samples in a bid to avoid potential contact with body fluids that could expose them to COVID-19 infection. Additionally, providers in the during-COVID-19 sample were more likely to avoid prescribing inappropriate medication (antibiotics, anti-TB medication, and steroids). This is a positive result that may reflect the efforts of the Nigerian government, SHOPS Plus, and other relevant actors to increase antimicrobial stewardship efforts within the private health system (41–43). More specifically, based on the results of the 2019 SP study which showed significant gaps in treatment initiation, SHOPS Plus implemented targeted interventions including trainings and webinars to improve the counseling and treatment initiation skills of networked providers and developed Information, Education and Communication (IEC) materials for patients and job aids for providers to support these efforts before the 2021 SP survey.

Our overall findings agree with those found among private providers in similar contexts (9,12,44,45). Few providers in our study prescribed TB medications, steroids, or other antibiotics, particularly among providers at SHOPS Plus supported clinical facilities (13%) and non-network clinical facilities (9%). This compared favorably with reports from other studies of clinical provider behavior, including 55% of SPs being prescribed antibiotics in Kenya (46), 61% in China (47), 68% in urban Indonesia (28), 76% of cases in South Africa (45), and in over 80% of cases in India (9,10,44).

### Limitations

Our study has several limitations. As with other SP studies, these results reflect a single interaction with a provider at one point in time and do not reflect situations where patients would have multiple visits to the same provider. Additionally, the naïve presumptive TB case used in this study only simulates the initial care-seeking step of a patient’s TB care journey rather than the longer TB case management protocol covering initial care-seeking, diagnosis, and treatment. This study employs the same methods as Rosapep, et al. (2022) which makes the two studies comparable, but due to its nature as a cross-sectional study, we cannot definitively state the cause of any changes observed between the 2019 and 2021 samples.

## Conclusion

Our study showed that few providers completed the benchmarked quality of care sequence, although they performed relatively well in the individual steps for history-taking and in refraining from prescribing unnecessary medications. Our results mirrored the 2019 study in most elements, except for a drop in clinical correctness among drug store providers. Overall, providers appeared mostly unconcerned about COVID-19 infection.

These results demonstrate that in the wake of the COVID-19 pandemic, and despite its limited impact in Nigeria, additional attention is still needed on the quality of TB services in the private sector in Nigeria to address the gap between the estimated number of new TB cases in Nigeria and the number of cases reported each year. Greater efforts to engage and support private providers, and implementing solutions such as working with drug shop proprietors to make referring for testing a standard part of their practice may help reduce the testing bottleneck at drug shops. Although Nigeria has maintained pre-pandemic levels of TB notification, it is important to establish high-quality screening by all providers to find the missing patients with TB and close the gap in TB notification.

## End Matter

### Data availability statement

The data that support the findings of this study are available from the corresponding author upon reasonable request.

### Author Contributions

A.S. was involved in study design, data collection, analysis, and interpretation and drafting of the manuscript. L.R., B.O.F., E.B., and B.J. were involved in study design, data collection, analysis, and interpretation, and critical revision of the manuscript. M.A.H.K. was involved in data analysis and interpretation. L.H. and N.A.V. were involved in data collection and critical revision of the manuscript. B.D. provided expert consultation on context and subject matter and critical manuscript revision. J.D., C.A., O.C-A., and M.P. were involved in study design, provided expert consultation on context and subject matter, and critical manuscript revision. C.O-O. was involved in study design, data collection, analysis, and interpretation, provided expert consultation on context and subject matter, and critical manuscript revision. All contributing authors have read and approved the manuscript prior to submission.

### Ethical approval

Ethics committee of the McGill University Health Centre gave ethical approval for this work (COVID BMGF / 2021-7197). IRB of Georgetown University gave ethical approval for this work (Georgetown-MedStar IRB System STUDY00003422). Ethics committee of the Health Research Ethics Committee Lagos State University Teaching Hospital gave ethical approval for this work (LREC/06/10/1517). Ethics committee of the Health Research Ethics Committee Kano State Ministry of Health gave ethical approval for this work (MOH/Off797/T.I/2168).

### Funding

This work was supported by the Bill & Melinda Gates Foundation [grant number INV-022420] and the SHOPS Plus program funded by USAID [Contract #: AID-OAA-A-15-00067]. The funders had no role in the study design, data collection, data analysis, decision to publish, or writing of the manuscript.

## Acknowledgements

The authors would like to thank the COVET study team and the McGill International TB Centre for sustained support throughout this study, particularly Caroline Vadnais and Rishav Das. We are indebted to the hard work from the entire SHOPS Plus Nigeria team, particularly Micah Sorum and Modupe Toriola; Mariose Amarikwa and Qualiquant Services, LTD, who trained and supervised the standardized patients in this study; and the standardized patient actors who conducted the health facility visits in this study. The authors also thank Guy Stallworthy of the Bill & Melinda Gates Foundation for his support.

