## Supplement 1: Additional methods for "Private sector tuberculosis care quality during the COVID-19 pandemic: A repeated cross-sectional standardized patients study of adherence to national TB guidelines in urban Nigeria"

**Standardized Patient recruitment and training**

All staff contracted to serve as SPs and supervisors for the 2021 survey were part of the 2019 survey. Prior to selection for the 2021 survey, staff were interviewed for suitability and availability and were also subject to a health check to ensure they did not have any underlying health conditions which would interfere with their ability to present the SP case. The Lagos team, which consisted of 12 staff (seven SPs, three Junior Supervisors, and two Senior Supervisors), and the Kano team of nine staff (six SPs, two Junior Supervisors, and one Senior Supervisor) were invited to a centralized training workshop. The training took place in Lagos for all members of the study team from Kano and Lagos from 19-30 April 2021. The first week consisted of a six-day, 48-hour training including extensive discussions and role plays. Study materials were adapted from the 2019 survey with few modifications (Kwan A. , et al., 2019; Rosapep, et al., 2022). Training participants scrutinized the scenario scripts to ensure they were appropriate for the local context. During the training, SPs conducted several rounds of full mock interviews to help them immerse in their roles and avoid pitfalls that could compromise the study. The training also emphasized mitigation protocols and techniques to protect SPs from harm (i.e., avoiding invasive procedures such as blood draws, administration of medicines, etc.).

**Survey piloting**

In the second week of training, all SPs conducted dry runs in eight facilities. The facilities visited were the same dry run facilities used in the 2019 survey. Immediately after, there was a full day debriefing session to discuss the field encounters, challenges, possible mitigation strategies and learnings. Following the dry run, the Kano team returned to their home state to conduct a full pilot exercise for the remainder of the week while the Lagos team carried on with their pilot in their state. The piloting was intended to provide additional SP practice and to test logistics and feasibility of the intended daily fieldwork and administration of the exit interview in various field settings. In Lagos, SPs completed pilot visits in 23 facilities; in Kano SPs completed visits in 16 facilities. All pilot facilities were previously used as pilot facilities in 2019 and were not part of the 2021 sample frame.
